# Changes in Treatment and Severity of Multisystem Inflammatory Syndrome in Children: An EHR-based cohort study from the RECOVER program

**DOI:** 10.1101/2022.10.19.22281256

**Authors:** Julia Schuchard, Deepika Thacker, Ryan Webb, Charles Bailey, Tellen D. Bennett, Jonathan D. Cogen, Ravi Jhaveri, Pei-Ni Jone, Grace M. Lee, Mitchell Maltenfort, Asuncion Mejias, Colin M. Rogerson, Grant S. Schulert, Eneida A. Mendonca, the RECOVER consortium

**Author notes:** **Address correspondence to:** Julia Schuchard, 2716 South St., Philadelphia, PA 19146. Contributed equally as co-first authors (Julia Schuchard, Deepika Thacker). **Authorship:** Authorship has been determined according to ICMJE recommendations. **Role of Funder/Sponsor:** The NIH had no role in the design and conduct of the study. The content is solely the responsibility of the authors and does not necessarily represent the official views of the RECOVER Program, the NIH or other funders. **Data Sharing Statement:** Deidentified individual participant data will not be made available.

## Abstract

**Objectives:** The purpose of this study was to examine how the treatment and severity of multisystem inflammatory syndrome in children (MIS-C) has changed over more than two years of the COVID-19 pandemic in the United States.

**Methods:** Electronic health record data were retrieved from the PEDSnet network as part of the NIH Researching COVID to Enhance Recovery (RECOVER) Initiative. The study included data for children ages 0 to 20 years hospitalized for MIS-C from March 1, 2020 through July 20, 2022. Descriptive statistics for MIS-C treatments and laboratory results were computed for three time periods of interest: March 1, 2020 – May 31, 2021 (pre-Delta); June 1 – December 31, 2021 (primarily Delta); January 1 – July 20, 2022 (primarily Omicron). Standardized differences measured the effect size of the difference between Omicron and pre-Omicron cohorts.

**Results:** The study included 946 children with a diagnosis of MIS-C. The largest differences in the Omicron period compared to prior years were decreases in the percentage of children with abnormal troponin (effect size = 0.40), abnormal lymphocytes (effect size = 0.33), and intensive care unit (ICU) visits (effect size = 0.34). There were small decreases in the Omicron period for the majority of treatments and abnormal laboratory measurements examined, including infliximab, anticoagulants, furosemide, aspirin, IVIG without steroids, echocardiograms, mechanical ventilation, platelets, ferritin, and sodium.

**Conclusions:** This study provides the first evidence that the severity of MIS-C declined in the first half of the year 2022 relative to prior years of the COVID-19 pandemic in the United States.

**Article Summary:** Using electronic health record data for 946 children, we found evidence that the severity of MIS-C declined during the first half of the year 2022.

**What’s Known on This Subject:** The clinical management of multisystem inflammatory syndrome in children (MIS-C) has commonly included intravenous immune globulin, steroids, and non-steroidal anti-inflammatory agents. Many children with MIS-C have required intravenous fluids, inotropes and vasopressors, and in some cases, mechanical ventilation.

**What This Study Adds:** Recent decreases in the percentage of children with MIS-C that have abnormal troponin, abnormal lymphocytes, or intensive care unit visits provide evidence that the severity of MIS-C has declined in the first half of the year 2022.

## Introduction

In the spring of 2020, physicians in Europe alerted the world to an inflammatory condition in children resembling Kawasaki disease and suggested a likely correlation with SARS-CoV-2 infection.^1-3^ In May 2020, the Centers for Disease Control and Prevention (CDC) outlined a set of diagnostic criteria for this condition that was termed multisystem inflammatory syndrome in children associated with COVID-19 (MIS-C).^4^ COVID cases are defined as per the CDC guidance (see https://ndc.services.cdc.gov/conditions/coronavirus-disease-2019-2021). MIS-C is now considered one of the pediatric Post-Acute Sequelae of SARS-CoV-2 infection (PASC) conditions, defined as ongoing, relapsing, or new symptoms or other health effects occurring four or more weeks after acute SARS-CoV-2 infection.

Due to its resemblance to Kawasaki disease, initial treatment protocols applied a similar management with intravenous immune globulin (IVIG) and non-steroidal anti-inflammatory agents.^1^ In July 2020 the American College of Rheumatology released recommendations that included IVIG and aspirin, with glucocorticoids or anakinra administration in more severe cases^5^ Recommendations were subsequently updated and included initial use of other immunomodulatory treatment.^6^ The most recent version (Version 3) recommends IVIG and steroids rather than IVIG alone due to high failure rates in cohort studies.^7^ Unlike most children with Kawasaki Disease, many children presenting with MIS-C required supportive care in the form of intravenous fluids, inotropes and vasopressors, and in some cases, mechanical ventilation.^8,9^ An analysis of treatments for MIS-C in the United States from February 2020 to July 2021 found that the most common treatments were IVIG, steroids, and antiplatelet medications, and the use of those treatments increased over time.^10^

Changes in the clinical management of MIS-C are likely to occur as new knowledge about the condition emerges, clinicians gain more experience treating children with MIS-C, the clinical presentation of MIS-C changes over time, and institutional guidelines change. The purpose of this study was to examine how the treatment and severity of MIS-C has differed during the recent Omicron period (beginning early in 2022) compared to prior phases of the pandemic in the United States using a large electronic health record (EHR) database. Although pediatric cases of SARS-CoV-2 infection increased dramatically with the rise of the Omicron variant, studies show recent declines in the rate of MIS-C relative to confirmed SARS-CoV-2 infections.^11-14^ We hypothesized that the severity of MIS-C has also decreased in the Omicron period, which would be indicated by decreases in the percentage of children with abnormal laboratory measurements as well as intensive supportive treatments.

## Methods

This study is part of the NIH Researching COVID to Enhance Recovery (RECOVER) Initiative, which seeks to understand, treat, and prevent PASC (for more information on RECOVER, visit recovercovid.org). Data for this study were drawn from PEDSnet (pedsnet.org), a national network of children’s health systems that share EHR data to conduct research and improve child health. The study included data from March 1, 2020 through July 20, 2022. Participating institutions included Children’s Hospital of Philadelphia, Cincinnati Children’s Hospital Medical Center, Children’s Hospital Colorado, Ann & Robert H. Lurie Children’s Hospital of Chicago, Nationwide Children’s Hospital, Nemours Children’s Health System (a Delaware and Florida health system), Seattle Children’s Hospital, and Stanford Children’s Health. The Children’s Hospital of Philadelphia’s institutional review board designated this study as not human subjects research and waived the need for consent.

Children with MIS-C were identified by clinicians’ diagnosis, as recorded in the electronic health records. We identified MIS-C diagnoses using the MIS-C ICD-10CM code (M35.81) and a string searching method to look for matches in the diagnosis term from the proprietary point-of-care terminology (Intelligent Medical Objects (Rosemont, IL)) used at all participating institutions. A match is made via string searching when the diagnosis term contains the token ‘MIS-C’ or when both the token ‘multisystem’ and the subword token ‘inflam’ are present in the same term. The cohort entrance date was defined as the date of the patient’s first recorded diagnosis of MIS-C in the database. Children in the study were less than 21 years old at cohort entry and had an inpatient visit associated with the MIS-C diagnosis. To exclude children who may have been treated with IVIG or systemic steroids for pre-existing conditions, we excluded children with any recorded exposure to these medications prior to the 30 days before the MIS-C diagnosis (n = 253 excluded).

Medications, procedures, and laboratory measurements in the study were selected based on literature review, consensus among expert clinicians on the study team, and data quality (e.g., low rates of missingness in the study cohort). Of the eight laboratory measurements included in the study, troponin, platelets, lymphocytes, and creatinine were considered the most relevant because they have been identified as important indicators of MIS-C severity.^15,16^

Descriptive statistics for the variables of interest were computed using R 4.1.2^17^ for the cohort overall and separately according to three time periods of cohort entry based on the emergence of coronavirus variants in the United States:^18^ March 1, 2020 – May 31, 2021 (pre-Delta); June 1 – December 31, 2021 (primarily Delta); January 1 – July 20, 2022 (primarily Omicron). We calculated the percentage of children that received each treatment at any time from 7 days prior to 30 days after the MIS-C diagnosis. We also calculated the percentage of children with an intensive care unit (ICU) visit within 7 days before or after the diagnosis. For each laboratory variable, we calculated the number of children with at least one abnormal value versus only normal values during the first 3 days of hospitalization. The following cut-points were used to define abnormal laboratory values: platelets < 150,000/microliter, lymphocytes < 4,500/microliter if age < 1 year or < 1,500/microliter if age ≥ 1 year, serum/plasma troponin > 0.1 ng/mL, creatinine > 0.7 mg/dL if age < 3 years or > 1.0 mg/dL if age ≥ 3 years, C-reactive protein ≥ 3 mg/dL, erythrocyte sedimentation rate ≥ 40 mm/hr, ferritin > 200 ng/mL, and sodium < 135 mmol/L.

We then collapsed the data for each variable of interest across children in the Delta and pre-Delta time periods and compared them to children in the Omicron period. In addition, we calculated standardized differences to measure the effect size of the difference between Omicron and pre-Omicron cohorts. Standardized differences, which are often used to characterize the balance between groups in propensity-adjusted studies, are useful to measure effect sizes when comparing categorical variables, just as Cohen’s d may be used when comparing continuous variables (e.g., 0.5 = one-half the standard deviation of the variable). To examine the distributions of laboratory values in the Omicron versus pre-Omicron time periods, we plotted the distributions of values using the highest (for troponin, creatinine, C-reactive protein, erythrocyte sedimentation rate, and ferritin) or lowest (for platelets, lymphocytes, and sodium) value per person during the first three days of hospitalization.

## Results

From March 1^st^ 2020 through July 20^th^ 2022, we identified 946 children with a diagnosis of MIS-C. Child characteristics overall and in each time period of interest are reported in Table 1. The median age was 8 years, and 60% were male, 21% Hispanic, 41% non-Hispanic white, 23% non-Hispanic Black, and 15% of multiple or other race/ethnicities. In the Omicron period, over a third (37%) of children with MIS-C were less than 5 years old compared to 29% of children prior to 2022.

**Table 1.**
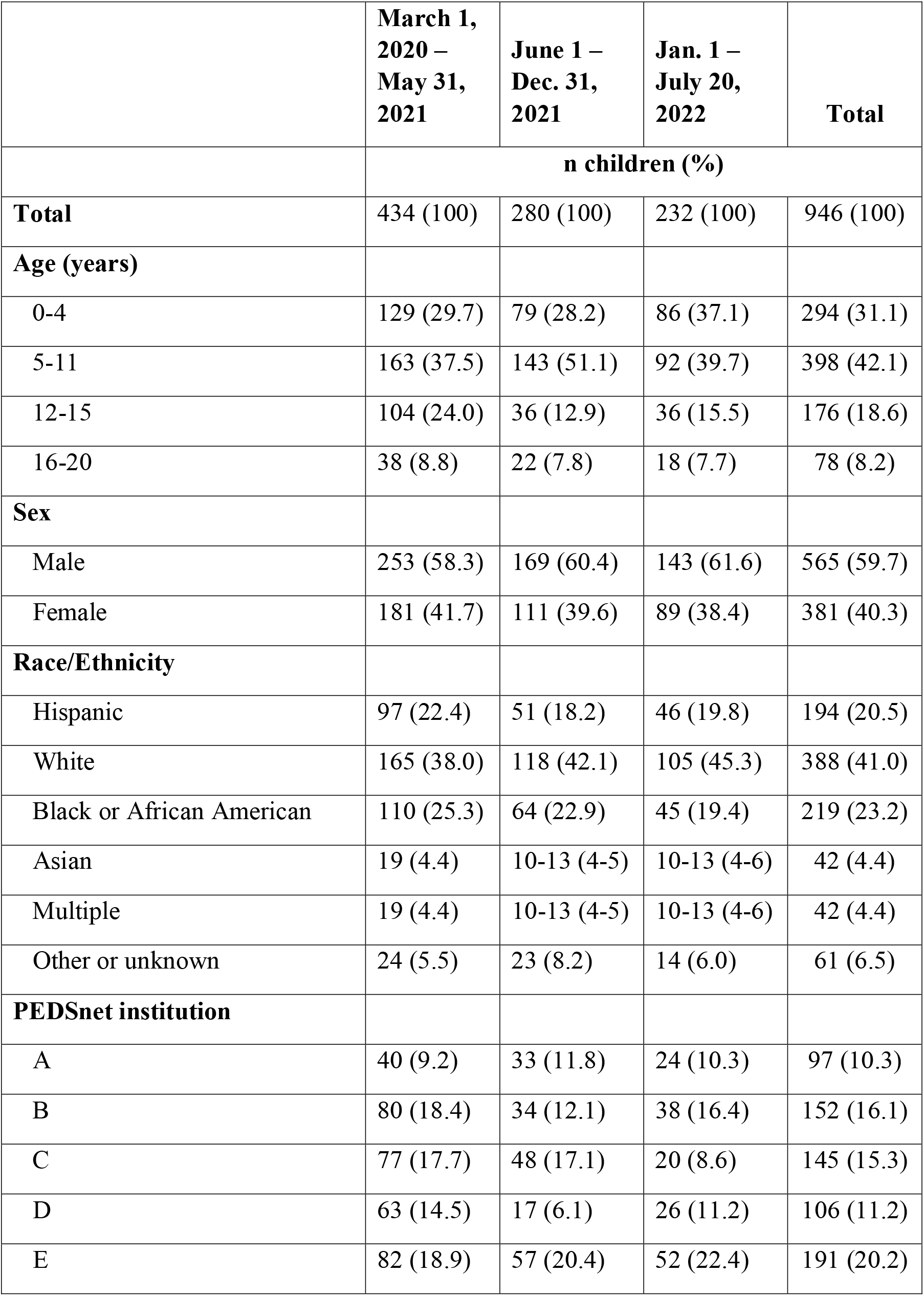
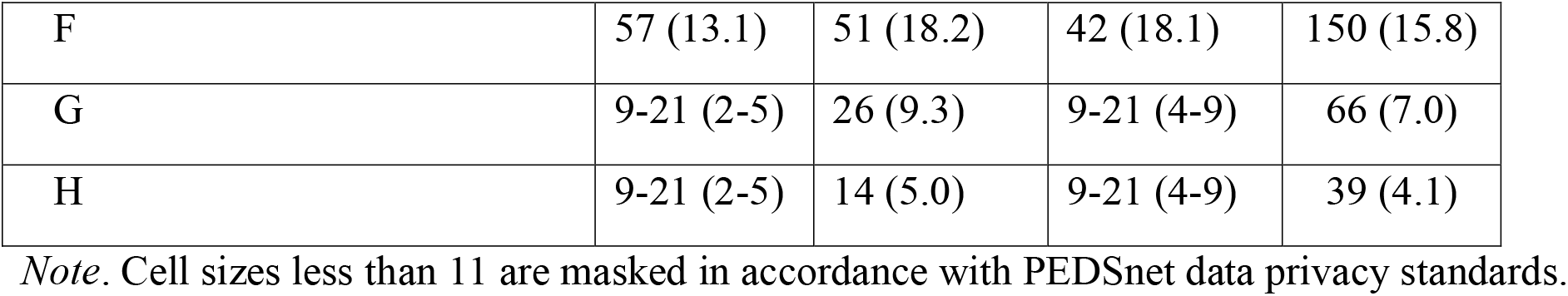
Patient Characteristics

|  | **March 1,**<br>**2020 –**<br>**May 31,**<br>**2021** | **June 1 –**<br>**Dec. 31,**<br>**2021** | **Jan. 1 –**<br>**July 20,**<br>**2022** | **Total** |
| --- | --- | --- | --- | --- |
|  | **n children (%)** |  |  |  |
| **Total** | 434 (100) | 280 (100) | 232 (100) | 946 (100) |
| **Age (years)** |  |  |  |  |
| 0-4 | 129 (29.7) | 79 (28.2) | 86 (37.1) | 294 (31.1) |
| 5-11 | 163 (37.5) | 143 (51.1) | 92 (39.7) | 398 (42.1) |
| 12-15 | 104 (24.0) | 36 (12.9) | 36 (15.5) | 176 (18.6) |
| 16-20 | 38 (8.8) | 22 (7.8) | 18 (7.7) | 78 (8.2) |
| **Sex** |  |  |  |  |
| Male | 253 (58.3) | 169 (60.4) | 143 (61.6) | 565 (59.7) |
| Female | 181 (41.7) | 111 (39.6) | 89 (38.4) | 381 (40.3) |
| **Race/Ethnicity** |  |  |  |  |
| Hispanic | 97 (22.4) | 51 (18.2) | 46 (19.8) | 194 (20.5) |
| White | 165 (38.0) | 118 (42.1) | 105 (45.3) | 388 (41.0) |
| Black or African American | 110 (25.3) | 64 (22.9) | 45 (19.4) | 219 (23.2) |
| Asian | 19 (4.4) | 10-13 (4-5) | 10-13 (4-6) | 42 (4.4) |
| Multiple | 19 (4.4) | 10-13 (4-5) | 10-13 (4-6) | 42 (4.4) |
| Other or unknown | 24 (5.5) | 23 (8.2) | 14 (6.0) | 61 (6.5) |
| **PEDSnet institution** |  |  |  |  |
| A | 40 (9.2) | 33 (11.8) | 24 (10.3) | 97 (10.3) |
| B | 80 (18.4) | 34 (12.1) | 38 (16.4) | 152 (16.1) |
| C | 77 (17.7) | 48 (17.1) | 20 (8.6) | 145 (15.3) |
| D | 63 (14.5) | 17 (6.1) | 26 (11.2) | 106 (11.2) |
| E | 82 (18.9) | 57 (20.4) | 52 (22.4) | 191 (20.2) |
| F | 57 (13.1) | 51 (18.2) | 42 (18.1) | 150 (15.8) |
| G | 9-21 (2-5) | 26 (9.3) | 9-21 (4-9) | 66 (7.0) |
| H | 9-21 (2-5) | 14 (5.0) | 9-21 (4-9) | 39 (4.1) |

*Note.* Cell sizes less than 11 are masked in accordance with PEDSnet data privacy standards.

MIS-C treatments overall and in each time period are reported in Table 2. The most commonly used medications were aspirin (73% of children) and a combination of IVIG and steroids (57%). Echocardiograms were commonly performed (90%). The following MIS-C treatments were excluded from Table 2 because they were used for only 1-10 children in the cohort (≤ 1%): remdesivir (an antiviral agent), tocilizumab (an interleukin-6 anatagonist), enalapril, dialysis, and extracorporeal membrane oxygenation (ECMO). The use of infliximab and anakinra was also low, with <15% of children receiving each of these medications, and the use of infliximab was almost exclusively observed in just one of the eight participating institutions.

**Table 2.** Medications and Procedures

|  | **March 1,**<br>**2020 –**<br>**May 31,**<br>**2021** | **June 1 –**<br>**Dec. 31,**<br>**2021** | **Jan. 1 –**<br>**July 20,**<br>**2022** | **Total** | **March 1,**<br>**2020 – Dec.**<br>**31, 2021 vs**<br>**Jan. 1 – July**<br>**20, 2022**<br>**(standardized**<br>**difference)** |
| --- | --- | --- | --- | --- | --- |
|  | **n children (%)** |  |  |  |  |
| **Total** | 434 (100) | 280<br>(100) | 232<br>(100) | 946 (100) |  |
| Aspirin | 343 (79.0) | 192<br>(68.6) | 151<br>(65.1) | 686 (72.5) | 0.22 |
| IVIG and steroids | 268 (61.8) | 141<br>(50.4) | 130<br>(56.0) | 539 (57.0) | 0.03 |
| IVIG (no steroids) | 69 (15.9) | 52<br>(18.6) | 23 (9.9) | 144 (15.2) | 0.21 |
| Steroids (no IVIG) | 41 (9.4) | 45<br>(16.1) | 24 (10.3) | 110 (11.6) | 0.05 |
| IV fluid resuscitation | 382 (88.0) | 246<br>(87.9) | 197<br>(84.9) | 825 (87.2) | 0.09 |
| Vasoactive medications | 135 (31.1) | 88<br>(31.4) | 66 (28.5) | 289 (30.6) | 0.06 |
| Infliximab | 65 (15.0) | 46<br>(16.4) | 15 (6.5) | 126 (13.3) | 0.29 |
| Anakinra | 42 (9.7) | 23 (8.2) | 17 (7.3) | 82 (8.7) | 0.07 |
| Anticoagulants | 176 (40.6) | 97<br>(34.6) | 73 (31.5) | 346 (36.6) | 0.14 |
| Furosemide | 141 (32.5) | 79<br>(28.2) | 53 (22.8) | 273 (28.9) | 0.18 |
| Echocardiogram | 408 (94.0) | 244<br>(87.1) | 196<br>(84.5) | 848 (89.6) | 0.21 |
| Mechanical ventilation | 82 (18.9) | 52<br>(18.6) | 28 (12.1) | 162 (17.1) | 0.19 |
| ICU visit | 203 (46.8) | 113 (40.4) | 65 (28.0) | 381 (40.3) | 0.34 |
| **Median per patient (interquartile range)** |  |  |  |  |  |
| Length of inpatient stay (days) | 5 (4-7) | 5 (3.5 – 7) | 5 (3 – 6.5) | 5 (4 – 7) | 0.13 |
| Number of echocardiograms during inpatient stay | 4.5 (2 – 9) | 3 (2 – 8) | 3 (1-7) | 4 (2 – 8) | 0.20 |

*Note.* The codeset used to search for steroids included intravenous and oral prednisone, prednisolone, methylprednisone, and dexamethasone. The IV fluids codeset included sodium chloride or a combination of calcium chloride, potassium chloride, sodium chloride, and sodium lactate. The vasoactive medication codeset included vasopressin, dobutamine, milrinone, dopamine, dobutamine, lidocaine, epinephrine, norepinephrine, phenylephrine, isoproterenol. The anticoagulants codeset included heparin, low molecular weight heparin, enoxaparin, and dalteparin.

MIS-C laboratory measurements overall and in each time period are reported in Table 3. Distributions for each variable are depicted in Figure 1. Of the eight measurements included in the study, troponin showed the largest difference in the Omicron period compared to prior periods, both in the percentage of patients with at least one abnormal value (see Table 3) and the distribution of the highest value per person (see Figure 1).

**Table 3.** Laboratory Measurements

|  | **March 1, 2020 – May 31, 2021** | **June 1 – Dec. 31, 2021** | **Jan. 1 – July 20, 2022** | **Total** | **March 1, 2020 – Dec. 31, 2021 vs Jan. 1 – July 20, 2022 (standardized difference)** |
| --- | --- | --- | --- | --- | --- |
| **Platelets** |  |  |  |  |  |
| Has measurement: n | 263 | 190 | 157 | 610 |  |
| Abnormal < 150,000/microliter: n (%)* | 127 (48.3) | 82 (43.2) | 59 (37.6) | 268 (43.9) | 0.17 |
| **Lymphocytes** |  |  |  |  |  |
| Has measurement: n | 313 | 226 | 195 | 734 |  |
| Abnormal (age < 1, < 4,500/microliter OR age ≥ 1, < 1,500/microliter): n (%) | 251 (80.2) | 167 (73.9) | 122 (62.6) | 540 (73.6) | 0.33 |
| **Troponin** |  |  |  |  |  |
| Has measurement: n | 344 | 191 | 146 | 681 |  |
| Abnormal (> 0.1 ng/mL): n (%) | 175 (50.9) | 68 (35.6) | 39 (26.7) | 282 (41.4) | 0.40 |
| **Creatinine** |  |  |  |  |  |
| Has measurement: n | 421 | 272 | 227 | 920 |  |
| Abnormal (age < 3yr, > 0.7 mg/dL OR age ≥ 3, > 1.0 mg/dL): n (%) | 50 (11.9) | 35 (12.9) | 23 (10.1) | 108 (11.7) | 0.07 |
| **C-reactive protein** |  |  |  |  |  |
| Has measurement: n | 427 | 268 | 226 | 921 |  |
| Abnormal (≥ 3 mg/dL): n (%) | 405 (94.9) | 252 (94.0) | 215 (95.1) | 872 (94.7) | 0.03 |
| **Erythrocyte sedimentation rate** |  |  |  |  |  |
| Has measurement: n | 380 | 240 | 201 | 821 |  |
| Abnormal ($\geq 40$ mm/hr): n (%) | 255<br>(67.1) | 149<br>(62.1) | 126<br>(62.7) | 530<br>(64.6) | 0.05 |
| **Ferritin** |  |  |  |  |  |
| Has measurement: n | 389 | 234 | 187 | 810 |  |
| Abnormal ($> 200$ ng/mL): n (%) | 297<br>(76.4) | 165<br>(70.5) | 117<br>(62.6) | 579<br>(71.5) | 0.25 |
| **Sodium** |  |  |  |  |  |
| Has measurement: n | 421 | 273 | 227 | 921 |  |
| Abnormal ($< 135$ mmol/L): n (%) | 294<br>(69.8) | 192<br>(70.3) | 144<br>(63.4) | 630<br>(68.4) | 0.14 |

\*The denominator is the number of children with at least one recorded measurement.

**Figure 1.**
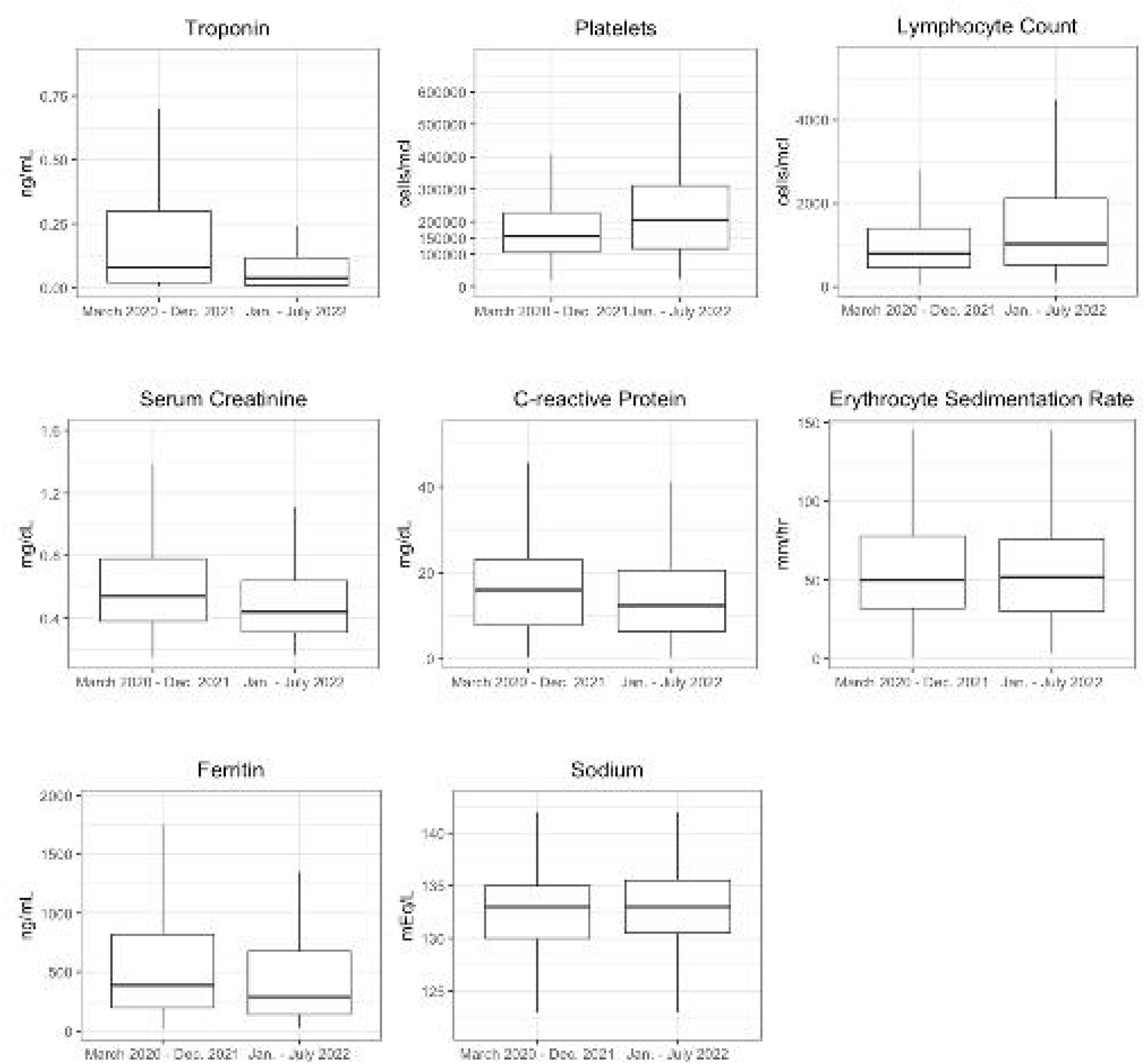
Boxplots depict the distributions of laboratory values in the Omicron period (Jan. – July 2022) compared to prior years (March 2020 – Dec. 2021). Each distribution includes the most abnormal value per person during the initial three days of hospitalization. Outliers (i.e., values outside 1.5 times the interquartile range above the upper quartile or below the lower quartile) were removed from the plots to improve clarity.

Of all the variables in this study, the largest differences in the Omicron period compared to prior years were decreases in the percentage of children with abnormal troponin (effect size = 0.40), abnormal lymphocytes (effect size = 0.33), and intensive care unit (ICU) visits, which decreased from 44% of children to 28% (effect size = 0.34). There were also small decreases in the Omicron period for many of the treatments and abnormal laboratory measurements we examined, with effect sizes between 0.1 and 0.3 for infliximab, anticoagulants, furosemide, aspirin, IVIG without steroids, echocardiograms, mechanical ventilation, platelets, ferritin, and sodium.

## Discussion

In this study of close to 1,000 children diagnosed with MIS-C in the United States, we examined medications, procedures, and laboratory testing associated with the clinical management of MIS-C over more than two years of the COVID-19 pandemic. As in prior studies,^10,19^ the most common treatments for MIS-C included IVIG, steroids, and aspirin, whereas the use of tocilizumab, remdesivir, dialysis, and ECMO were rare (<2% of children). For the majority of treatments that we examined in the study, there were small decreases in the percentage of children receiving medications or procedures during the Omicron period (January through mid-July 2022) compared to prior years (March 2020 – December 2021). Importantly, several findings indicate a decrease in the severity of MIS-C during the months of January through mid-July 2022 compared to prior years of the pandemic.

Factors contributing to the observed changes in treatment patterns may include earlier recognition of MIS-C cases leading to earlier initiation of first line therapies, a less severe MIS-C clinical phenotype associated with later SARS-CoV-2 variants, or changing institutional guidelines as new knowledge emerged. Recent recommendations to use IVIG and steroids (and not IVIG alone) for the treatment of MIS-C^7^ may have contributed to the observed decrease in the use of IVIG without steroids. The initial use of anticoagulation was likely related to high rates of thromboembolism reported in adults with COVID-19.^20,21^ As data were collected in children with MIS-C, analyses showed that the rate of thromboembolism was lower than adults with COVID,^22,23^ which may have led to the decrease in the use of anticoagulants observed in this study.

This study suggests a decrease in the severity of MIS-C during the months of January through mid-July 2022 compared to prior years of the pandemic. For three of the four laboratory results identified a priori as important indicators of MIS-C severity (i.e., troponin, platelets, and lymphocytes),^15,16^ the percentage of children with abnormal results declined across the three time periods examined in the study. These results suggest that the observed changes in treatment patterns were not solely attributable to evolving clinical guidelines or preferences for the management of MIS-C. The observed decrease in the use of intensive supportive management (i.e., ICU visits and ventilation) in this study also suggests a decrease in disease severity during the Omicron period. The results support other recent studies indicating a decline in the severity as well as the incidence of MIS-C.^12,13^, similar to findings that the acute severity of SARS-CoV-2 infection has decreased in the Omicron period for both children and adults.^24,25^

Demographic characteristics of the MIS-C cohort in this study showed a greater proportion of children under the age of 5 years in the Omicron period compared to prior phases. It is possible that COVID-19 vaccinations among older children have led to reduced incidence of SARS-CoV-2 infection and MIS-C relative to children less than 5 years, for whom vaccinations were only recently approved in the United States (June 2022). In addition, demographic data in this study corroborate prior findings that a relatively large proportion of children with MIS-C in the United States throughout the pandemic are Hispanic or Black, demonstrating racial and ethnic disparities in PASC.^25-28^

This study has limitations that highlight important future directions. We were unable to examine potential associations between vaccination and MIS-C treatment or severity because EHR documentation of COVID-19 vaccination had not yet been fully linked and harmonized with other EHR data in our network. As reliable vaccination data become available, it will be important to examine whether the observed decline in MIS-C severity and incidence may be attributable to increases in the proportion of the pediatric population vaccinated against COVID-19,^12,29^ the distinct clinical features of the Omicron variant,^30^ or both. Another important future direction is understanding the social determinants of health in relation to MIS-C and PASC. Although this and other studies have demonstrated racial and ethnic disparities in MIS-C, more research is needed to understand and mitigate the factors underlying these disparities. As geocoded environmental variables become available in the RECOVER network, there will be opportunities to examine factors such as local population density, income, education, employment, and housing. Finally, MIS-C diagnosis in this study was based on diagnoses in EHR rather than institutional registries of confirmed cases. This aspect of the study precludes the ability to confirm that all children with MIS-C met the CDC criteria for MIS-C,^4^ but it allows the capture of cases that may represent milder subphenotypes of MIS-C that do not meet all CDC criteria^16,31^ and/or cases that may not have been reported in institutional registries.

## Conclusions

This study provides the first evidence to our knowledge that the severity of MIS-C declined in the first half of the year 2022 relative to prior years of the COVID-19 pandemic in the United States. Among children diagnosed with MIS-C, there have been decreases in rates of elevated troponin and thrombocytopenia. The decline in abnormal troponin measurements is particularly important considering initial concerns about myocardial damage with MIS-C. The observed decline in many of the treatments used to manage MIS-C also has positive implications for clinical outcomes because there is presumably a lower risk of medication-related side effects. Continued monitoring of changes in the incidence, severity, and sociodemographic characteristics associated with MIS-C, as well as with acute SARS-CoV-2 infection among children, is important to inform standardized treatment guidelines and improve equitable care.

## Data Availability

Data will not be made available.

## Abbreviations

CDC: Centers for Disease Control and Prevention
EHR: electronic health record
ICU: intensive care unit
IVIG: intravenous immune globulin
MIS-C: multisystem inflammatory syndrome in children associated with COVID-19
PASC: Post-Acute Sequelae of SARS-CoV-2 infection

